# Whole Exome Sequencing Uncovers the Genetic Complexity of Bicuspid Aortic Valve in Families with Early Onset Complications

**DOI:** 10.1101/2024.02.07.24302406

**Authors:** Sara Mansoorshahi, Anji T Yetman, Malenka M Bissell, Yuli Y Kim, Hector Michelena, Dawn S Hui, Anthony Caffarelli, Maria G Andreassi, Ilenia Foffa, Dongchuan Guo, Rodolfo Citro, Margot De Marco, Justin T Tretter, Shaine A Morris, Simon C Body, Jessica X Chong, Michael J Bamshad, University of Washington Center for Rare Disease Research, BAVCon Investigators, EBAV Investigators, Dianna M Milewicz, Siddharth K Prakash

## Abstract

Bicuspid Aortic Valve (BAV) is the most common adult congenital heart lesion with an estimated population prevalence of 1%. We hypothesize that early onset complications of BAV (EBAV) are driven by specific impactful genetic variants. We analyzed whole exome sequences (WES) to identify rare coding variants that contribute to BAV disease in 215 EBAV families. Predicted pathogenic variants of causal genes were present in 111 EBAV families (51% of total), including genes that cause BAV (8%) or heritable thoracic aortic disease (HTAD, 17%). After appropriate filtration, we also identified 93 variants in 26 novel genes that are associated with autosomal dominant congenital heart phenotypes, including recurrent deleterious variation of *FBN2*, *MYH6*, channelopathy genes, and type 1 and 5 collagen genes. These findings confirm our hypothesis that unique rare genetic variants contribute to early onset complications of BAV disease.

## Introduction

Congenital heart disease (CHD) affects 8.2 per 1000 live births worldwide (1). Bicuspid Aortic Valve (BAV) is the most common adult CHD with an estimated population prevalence of 0.5-2%. BAV is a major cause of aortic stenosis or thoracic aortic aneurysms and is associated with other left ventricular outflow lesions such as coarctation (2). The robust heritability of BAV was demonstrated in studies showing that primary relatives are ten-fold more likely to have a BAV than matched controls (3). While more than 50% of individuals with BAV undergo aortic valve surgery during their lifetimes, phenotypic expression of BAV varies widely, with only approximately 10% of patients with BAV requiring aortic valve surgery in the first two decades of life (4,5).

We previously found that rare genomic copy number (CNV) variants involving congenital heart disease (CHD) genes are enriched in probands with early onset complications of BAV (EBAV), defined as clinical presentation due to aortic stenosis, aortic regurgitation, a large thoracic aortic aneurysm, or aortic dissection prior to age 30 (6). We hypothesized that this subset of individuals may have an increased burden of pathogenic genetic variants that drive aortic complications related to BAV disease. CNV analysis showed that genetic contributions to EBAV overlap with other congenital syndromes that include BAV such as Velocardiofacial syndrome and 1q21 deletion syndrome. We identified more than 50 large, potentially pathogenic CNVs in 10% of EBAV cases that overlap with CHD genes that can cause BAV that were not present in a cohort of older sporadic BAV cases. These observations also validate our hypothesis that the genetic contribution to EBAV is distinct compared to cases with later onset BAV complications.

We also compared the EBAV cohort to a cohort that was independently ascertained due to early onset thoracic aortic aneurysms or acute aortic dissections (ESTAD). More than 50% of ESTAD patients have BAV, but the contribution of known causal BAV genes to TAD is not known. To ascertain the impact of functionally annotated WES variants in the major BAV genes *GATA4*, *NOTCH1*, *SMAD6*, and *ROBO4,* we compared the prevalence of rare variants in these genes between cohorts and to controls without TAD. The burden of rare BAV gene variants was significantly enriched in EBAV cases compared to ESTAD cases, even when matched for the presence of BAV. These findings confirmed our hypothesis that the EBAV cohort is enriched for highly penetrant rare gene variants in known BAV genes, but that these genes do not significantly contribute to BAV in cohorts with TAD. Consistent with our earlier observations, we concluded that young patients who present primarily due to BAV are a genetically distinct subgroup with implications for genetic testing and prognosis. The overall goal of this study was to identify genetic variants that predispose to the EBAV phenotype.

## Methods

The study protocol was approved by the Committee for the Protection of Human Subjects at the University of Texas Health Science Center at Houston. All participants signed a written consent form, and study procedures were conducted in compliance with the ethical standards of the relevant national guidelines on human experimentation (HHS regulations 45 CFR part 46) and with the Helsinki Declaration of 1975, as revised in 2008.

Individuals were included in this study if they presented due to complications of BAV disease prior to age 30. Early onset complications were defined as greater than mild aortic stenosis or aortic regurgitation, a large thoracic aortic aneurysm (Z-score > 4), or aortic dissection. Individuals with known genetic mutations, genetic syndromes, or complex congenital heart disease were excluded.

The GenTAC cohort consists of whole exome sequencing (WES) data from 290 unrelated probands with BAV from the National Registry of Genetically Triggered Aortic Aneurysms and Other Cardiovascular Conditions (7). The JRRP cohort consists of WES data from 974 unrelated probands from the John Ritter Research Program in Aortic and Vascular Diseases at the University of Texas Health Science Center at Houston (8).

WES of the EBAV cohort was performed at the University of Washington Center for Rare Disease Research (UW-CRDR) using standard methods (see Supplemental Data). Variant filtration was performed using a customized version of seqr at the University of Washington Center for Rare Disease Research (v1.0-4cb7dfc3) with the following settings: de novo/dominant inheritance model; pathogenic, likely pathogenic, or VUS; nonsense, in frame, frameshift, or missense variants; REVEL > 0.5, CADD > 20, AF < 0.0001; pass variants only.

Missense variants were retained if they were predicted to be deleterious or disease-causing by at least 3 of polyphen, muttaster, SIFT, or FATHMM. Novel genes were prioritized if mutated in at least 3 unrelated EBAV probands. We filtered the final gene set to remove genes that cause recessive Mendelian phenotypes. The enrichment of rare variants was tabulated in 2x2 comparisons with selected deleterious rare variants in gnomAD version 2 data. gnomAD alleles were functionally annotated using FAVOR (https://favor.genohub.org).

## Results

### EBAV Cohort

We analyzed WES data from probands with early onset complication of bicuspid aortic valve disease (EBAV). The EBAV cohort consists of 215 families with 150 singletons, 16 duos, 34 trios, and 15 multiplex families. More than 95% of EBAV probands are from European ancestry populations. The mean age of EBAV probands at presentation was 18 years old, compared to the median age of 52 years in a recent meta-analysis of adult BAV cases (9)

### HTAD, BAV, and CHD gene variants in the EBAV Cohort

We first identified rare gene variants known to cause congenital heart disease (CHD) in the EBAV cohort. The list of candidate genes included 29 HTAD genes that are on current clinical sequencing panels, as well as 12 BAV genes and 190 CHD genes that have strong cumulative evidence to cause BAV or related congenital malformations from human or animal model data (Supplemental Data) (10,11).

Missense and loss of function variants predicted to be harmful by our analysis were selected using the criteria described in the Methods.

Predicted pathogenic variants of CHD genes were present in 111 (52%) EBAV families. In total, we discovered 138 alleles of 54 genes that cause dominant congenital heart phenotypes, primarily deleterious missense variants (75 probands). Predicted pathogenic variants of dominant CHD genes (including missense and LOF) comprised 77% of candidate variants, including 25 loss of function variants. We identified predicted pathogenic variants of BAV genes in 9 families (8%) and predicted pathogenic variants of HTAD genes in 19 families (17%). A total of 21 EBAV probands (19%) harbored deleterious rare variants in more than one candidate gene. Among all candidate genes, *MYH6* (6 missense, 1 LOF), *COL5A1* (5 missense), and *FBN2* (5 missense) harbored the most deleterious rare variants. The BAV genes with rare variants that passed all filters were *NOTCH1* (2 missense), *MIB1* (2 missense, 1 LOF) and *SMAD6* (2 missense, 2 LOF, 1 insertion). These observations suggest that genetic contributions to EBAV may differ significantly from the genetic landscape of later onset BAV disease.

### Novel Candidate Genes in the EBAV Cohort

Next, using the same criteria, we filtered the WES data to identify genes with rare deleterious variants in at least three unrelated EBAV probands (Figure 2).

**Figure 1:**
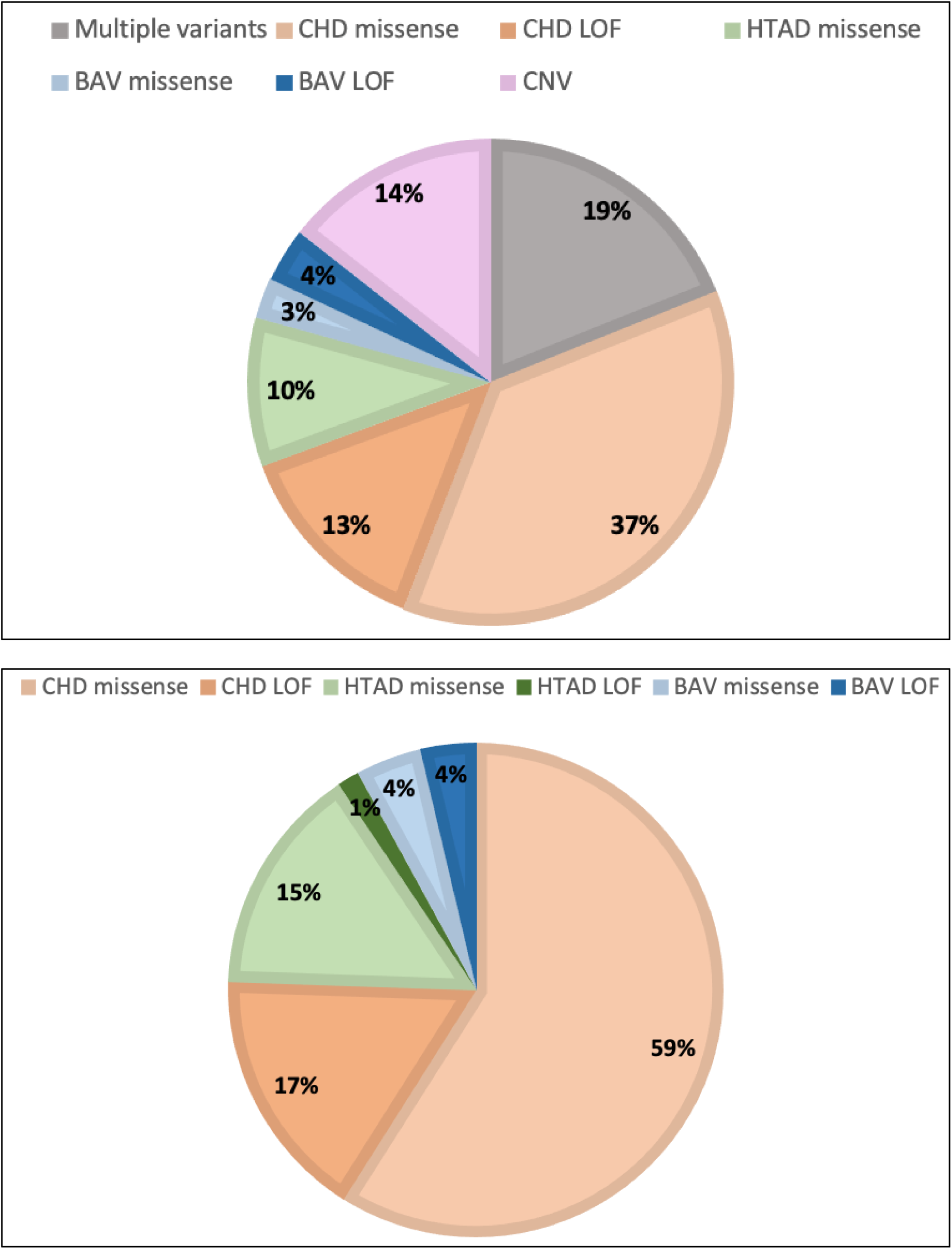
Distribution of predicted deleterious variants in 111 EBAV families and in candidate genes. **A.** Distribution of variants in EBAV families; **B.** Distribution of variants in candidate genes.

**Figure 2:**
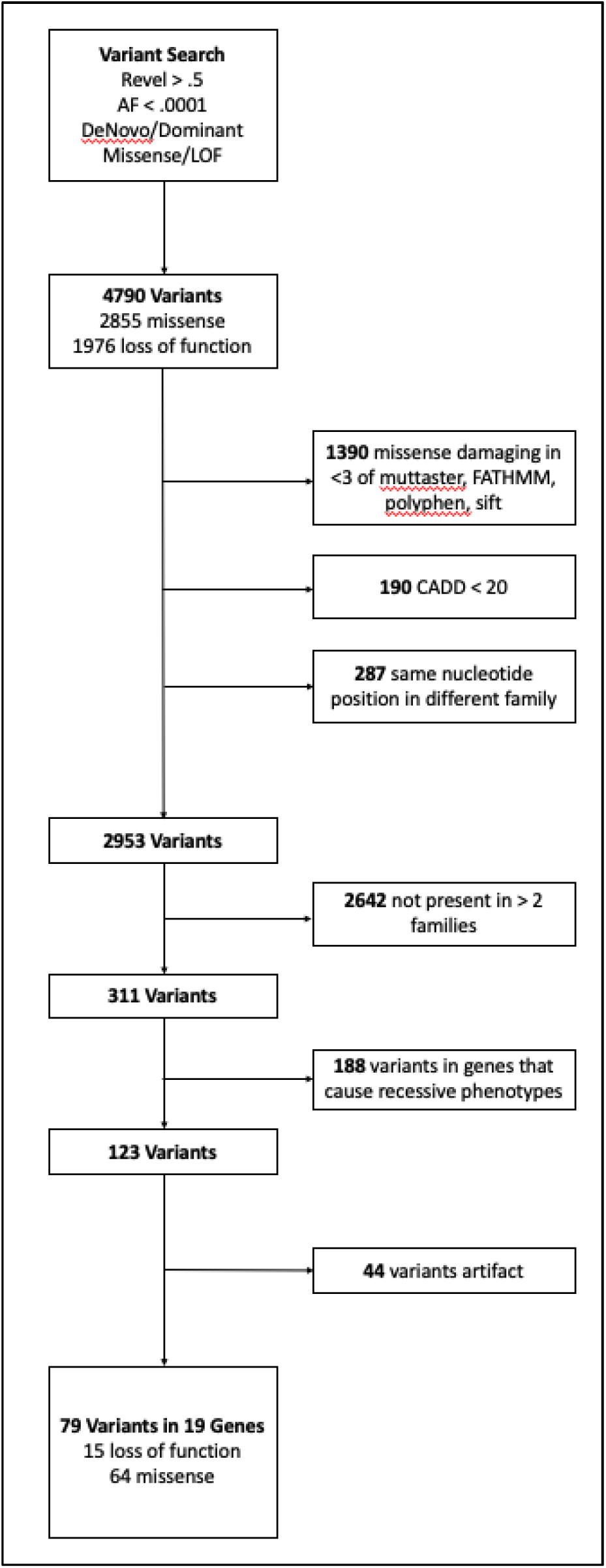
Filtration workflow to select candidate EBAV variants. The initial seqr project variant search generated 4790 candidates. Missense variants that were not rated as deleterious by at least three functional predictors, had normalized CADD scores < 20, and had the same base position in two different families were removed. We prioritized novel genes that had predicted deleterious variants in three or more families. We removed genes that are known to cause recessive or noncardiac phenotypes.

This selection strategy led to 79 variants in 19 genes associated with dominant cardiac phenotypes. The novel genes that are most highly enriched for deleterious rare variants are: *PKP2*, *MYH6*, *KCNH2, JAG1*, *KIF1A*, *TPTE2*, *COL1A1,* and *COL1A2* (Figure 3).

**Figure 3.**
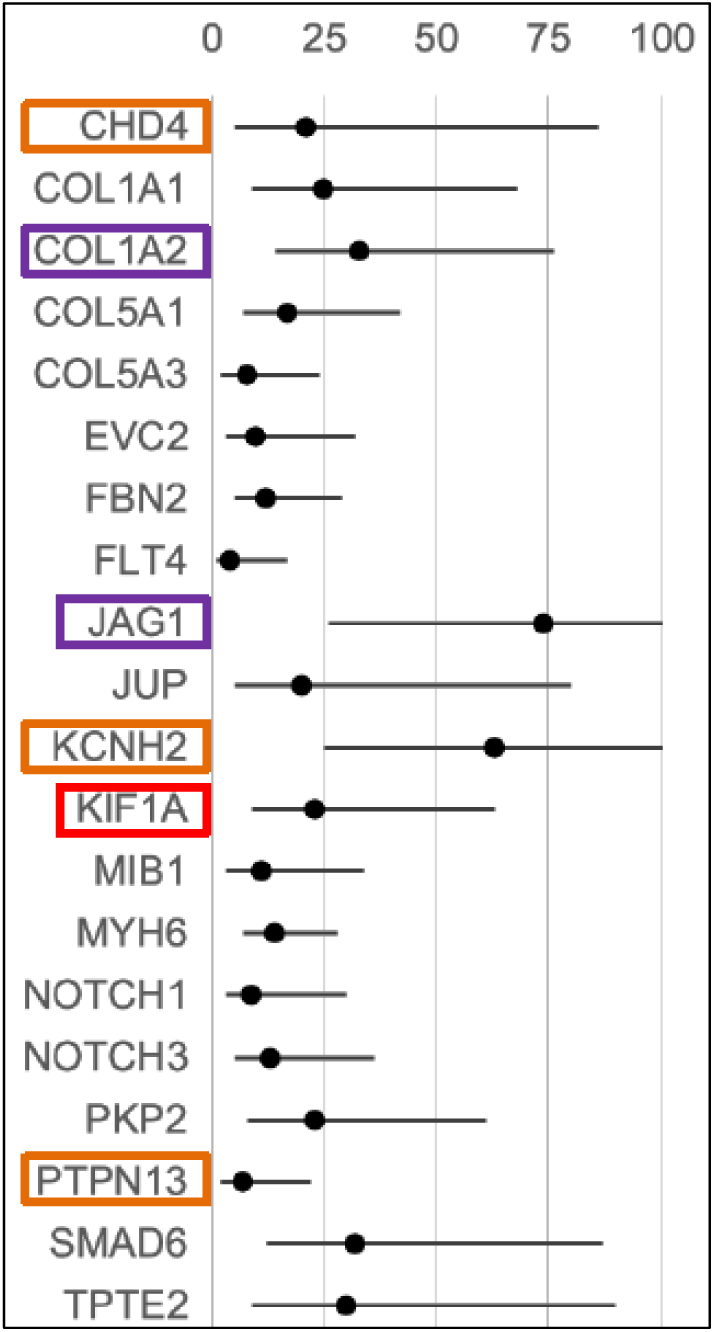
Enrichment of EBAV candidate gene variants. The number of rare deleterious loss of function and missense variants, selected according to the criteria defined in the Methods, were compared to the number of variants with equivalent functional classifications from European ancestry populations in gnomAD v2. Circles: enrichment vs. gnomAD (0-100X); horizontal lines: 95% confidence intervals; orange boxes: segregates with BAV in more than one EBAV family; purple boxes: also enriched in JRRP and GenTAC datasets; red boxes: at least one *de novo* variant.

Loss of function variants in *PKP2* were observed in three families. One rare variant that causes a stop-gain mutation (c.169C>T, p.Gln57Ter) is not present in gnomAD and has a CADD score of 41. A stop-gain mutation (c.235C>T, p.Arg79Ter, CADD 37, gnomAD frequency 4x10^-6) and a frameshift mutation (c.2198_2202del, p.His733ProfsTer8, CADD 29, gnomAD frequency 1.6x10^-5) are predicted to be pathogenic by ClinVar and are rare in gnomAD. The PKP2 frameshift variant did not segregate with BAV in one of the three EBAV families.

Six families harbored harmful missense or loss of function variants in *MYH6*. A single splice donor mutation c.1410+1G>A has not been reported in gnomAD and is predicted to be pathogenic by ClinVar. Three of the five missense variants are reported as variants of uncertain significance, and one missense variant has conflicting interpretations of pathogenicity. Notably, four of the 5 missense variants are not in the myosin motor or actin-binding domains (Supplementary Data). All five missense variants are reported to be damaging by MutTaster, FATHMM, and SIFT, with CADD scores > 22 and REVEL > 0.65. In one multiplex family with an *MYH6* variant, the variant is not present in two affected relatives (father and brother) and does not segregate with disease. The other families with an *MYH6* variant do not have additional samples available to determine segregation.

Rare deleterious *KCNH2* variants were observed in five families, including one loss of function variant and four missense variants. The loss of function frameshift variant c.33_34del (p.Ala13SerfsTer34) is not present in gnomAD and is predicted to be likely pathogenic by ClinVar. Three of four missense variants in *KCNH2* are not present in gnomAD, and the other missense variant of uncertain significance is rare in gnomAD (8x10^-6). All four missense variants have CADD > 20 and REVEL > 0.5. Three of the four missense variants are uniformly predicted to be damaging by Polyphen, SIFT, MutTaster, and FATHMM.

Rare Type 1 or Type 5 collagen gene variants were present in 21 families, primarily involving *COL1A1* (4), *COL1A2* (6), *COL5A1* (7), *COL5A2* (1) and *COL5A3* (4). All were missense variants except for a single *COL5A3* frameshift variant (c.2999del, p.Pro1000GlnfsTer144) that is present in gnomAD with a frequency of 3.18x10^-5 and has a CADD score of 27. The missense variants all had gnomAD frequencies < 1.5x10^-5 and are predicted to be damaging by Polyphen, SIFT, MutTaster, and FATHMM. In 6 out of 7 families with available segregation data, the missense variants were observed in apparently unaffected individuals (Supplemental Data). There were no other available samples to determine the segregation of the other missense or loss of function *COL1* or *COL5* variants.

Rare deleterious *KIF1A* variants were observed in four families, with two loss of function variants and two missense variants. The two predicted loss of function variants were a splice acceptor variant (c.4257-2A>C) and an in-frame deletion (c.718_720del, p.Lys240del). Both variants are not present in gnomAD and have CADD scores > 22. One missense variant (c.4222C>G, p.Leu1408Val) is also not present in gnomAD and has variable evidence for pathogenicity, with a REVEL score of 0.412 and a CADD score of 23.7. The second missense variant (c.1862C>T, p.Thr621Met) is present in gnomAD with a frequency of 1x10^-4^ and is reported as a variant of uncertain significance by ClinVar, but is predicted to be damaging by SIFT and MutTaster, with a CADD score of 22 and a REVEL score of 0.504. We determined that the splice acceptor variant is a *de novo* variant that is not present in two unaffected parents or in two unaffected siblings.

Microarray analysis also identified a heterozygous subtelomeric terminal deletion of 2q37.3 in 11 EBAV probands with a conserved breakpoint approximately 1 Mb telomeric to *KIF1A* (Supplemental Data). Given the distance between the breakpoint and the KIF1A promoter, we concluded that this recurrent 200 kilobase microdeletion is unlikely to affect *KIF1A* expression. No candidate genes with sequence variants that passed filtration are within the deleted subtelomeric region.

Several other gene variants were observed in at least three EBAV families, but there were no loss of function variants in these genes. There were *FBN2* missense variants in 5 families, and *FLNC*, *NOTCH3*, or *PTPN12* missense variants in 4 families.

### Developmental Functions of Candidate EBAV Genes

The GO processes that were most highly enriched in candidate EBAV genes that passed all filters were circulatory system development, actin filament-based movement, extracellular structure organization, multicellular organismal signaling, and cardiac development (Figure 4). The most highly enriched individual genes in these functional categories were *MYH6*, *KIF1A*, *JAG1, MIB1, PKP2*, and *SMAD6*, which all had predicted loss of function variants in the EBAV cohort. These data highlight the pleiotropic roles of cardiac developmental genes in aortic valve formation and cardiac function.

**Figure 4.**
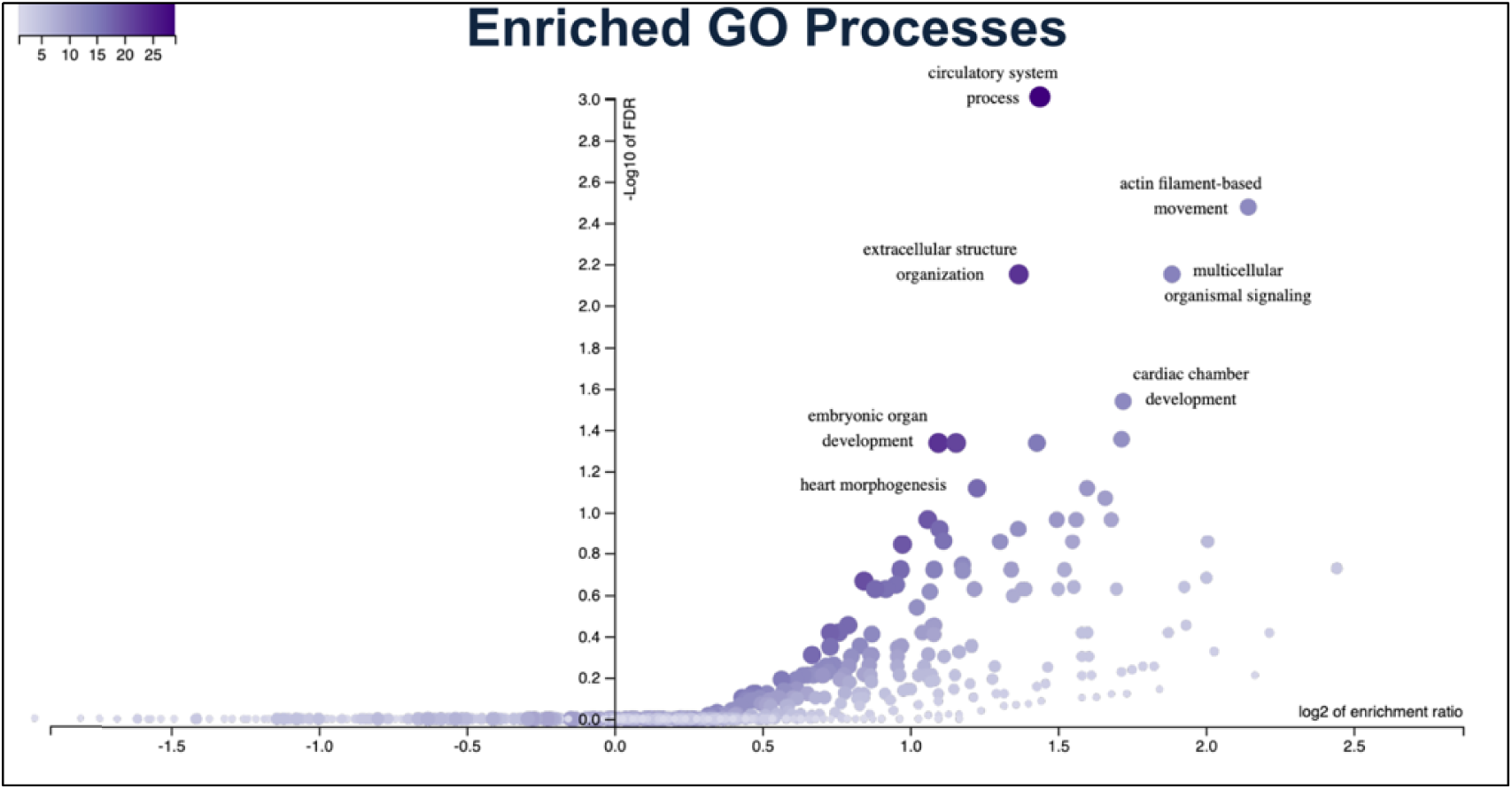
Gene ontology enrichment analysis of EBAV candidate genes. Plot was generated using Over-Representation Analysis of gene ontologies in WebGestalt 2019. The -log10 of false discovery rates (FDR) were plotted against the log2 of enrichment ratios.

### CHD Phenotypes of EBAV Cases with Rare Variants

In total, we identified at least one candidate gene mutation in 111 families (51%). Nineteen of these probands (17%) also presented with at least one other congenital lesion besides BAV. The most common congenital lesions were coarctation of the aorta in 12 families (63%) and mitral valve prolapse in two families. Seven probands presented with abnormal electrocardiographic findings (6%), primarily conduction delays ranging from first degree AV block to left bundle branch block. Several EBAV families with coarctation have variants in the same genes. Coarctation was observed in 2 of 5 families with variants in *COL1A1*, 2 of 4 families with variants in *FBN2*, and 2 of 4 families with variants in *MEGF8*. EBAV probands with rare sequence variants in candidate genes were not diagnosed at significantly younger ages and were not more likely to be diagnosed with thoracic aortic aneurysms or to undergo valve or aortic surgeries.

### Replication in TAD Datasets

Rare deleterious variants of several EBAV candidate genes are enriched in the EBAV cohort compared to gnomAD and are also present in two other TAD cohorts, JRRP and GenTAC: *COL1A2*, *COL5A1*, *FBN2*, *MIB1*, *MYH6*, and *NOTCH3* (Table 1). Rare deleterious variants of *JAG1*, *SMAD6*, *PKP2*, *EVC2*, *KCNH2*, and *KIF1A,* and *TPTE2* are also enriched in EBAV probands and are present in one other TAD cohort. Notably, we observed recurrent predicted loss of function variants of *MYH6*, *MIB1*, *JAG1*, *EVC2*, and *TPTE2*. *FLNC* harbored the largest number of rare variants but was not significantly enriched in EBAV probands. These observations suggest that genes that cause BAV may also contribute to TAD and are consistent with the increased prevalence of BAV in cohorts that present due to TAD.

**Table 1:** Rare variants of candidate genes in EBAV, GenTAC, and JRRP cohorts. *Indicates at least one loss of function allele in that dataset.

| Gene | EBAV (n=215) | GenTAC (n=200) | JRRP (n=974) |
| --- | --- | --- | --- |
| *NOTCH1* | 2 | 0 | 6 |
| *MIB1* | 3* | 3* | 6* |
| *JAG1* | 4 | 0 | 4* |
| *PKP2* | 3* | 1 | 0 |
| *MYH6* | 6* | 2* | 9 |
| *KCNH2* | 5* | 0 | 1 |
| *COL5A3* | 4* | 0 | 8 |
| *KIF1A* | 4* | 0 | 0 |
| *COL1A2* | 6 | 2 | 6 |
| *FBN2* | 5 | 1 | 12 |
| *FLNC* | 4 | 4 | 14 |
| *NOTCH3* | 4 | 3 | 4 |
| *SCN10A* | 3 | 8 | 3 |
| *SMAD6* | 5* | 2 | 0 |
| *COL1A1* | 4 | 2 | 3 |
| *COL5A1* | 7 | 2 | 8 |

### Segregation of Candidate Variants

Some candidate genes showed limited evidence for segregation in EBAV families (Supplemental Data). For example, *KCNH2* segregates in two duo families, and *PTPN13* segregates with BAV in one duo family and in one trio family. *CACHA1H*, encoding a voltage gated calcium channel subunit, segregated with BAV in a family with three affected individuals and one unaffected relative. *A de novo* variant of KIF1A was observed in a single family with 5 unaffected individuals (2 parents and 2 siblings) and 1 affected individual. Additionally, a *de novo* variant of *SCN8A* was observed in a single family with 3 unaffected individuals (2 parents and 1 sibling) and 1 affected individual. *CHD4*, *FBN2*, *NOTCH3*, and *JUP* segregated with BAV in single small families.

Other candidate genes showed evidence for reduced penetrance or lack of segregation. Notably, we observed that *SMAD6* missense and insertion alleles did not segregate with BAV in two families, although *SMAD6* loss of function alleles did segregate with BAV. *COL5A1* variants are present in two apparently unaffected individuals in separate families (50% of carriers). Similarly, only 3 of 7 *COL1A2* variant carriers (43%) in two families were diagnosed with BAV. The penetrance of *JAG1* (50%) was also reduced in two families with available data. In three trio families, *MYH6*, *COL5A3* and *PKP2* variants were all observed in one apparently unaffected parent. Incomplete segregation may suggest reduced penetrance of these variants or could imply that additional genetic mutations may be required to cause BAV in some families.

## Discussion

In this study, we analyzed WES data from 215 families (350 total participants) to identify genetic variants that cause early onset complications of BAV disease. We found that more than one-third of EBAV families have at least one variant in a likely pathogenic gene. In addition to known BAV and HTAD gene mutations, we observed recurrent deleterious variants in *MYH6*, *FBN2*, *KCNH2, PKP2*, type 1 and 5 collagens, and *KIF1A*.

We originally hypothesized that BAV and TAD may be caused by overlapping genes because BAV is enriched in some rare syndromic causes of TAD, and mutations in HTAD genes have been discovered in rare BAV cases with complex valvulo-aortopathy presentations (12,13) However, the results of this study reinforce our previous observations that there is minimal genetic overlap between young individuals who present primarily due to early onset valve complications and those who present due to early onset TAD (14). We did not identify any mutations of HTAD genes that cause Loeys-Dietz syndrome, which is associated with early onset TAD and BAV, and a total of three causal mutations in non-syndromic HTAD genes (a glycine substitution in FBN1 and loss of function mutations in SMAD4 and FOXE3). Correspondingly, we observed almost no thoracic aortic dissections in EBAV families, consistent with the low rates of aortic dissection in many BAV cohorts (15).

MYH6 encodes the alpha heavy-chain subunit of cardiac myosin. A recurrent missense mutation of MYH6 (p. Arg721Trp) was previously identified in 125 probands who had both coarctation of the aorta and BAV (16). In addition, MYH6 variants are implicated in a wide spectrum of congenital heart malformations, including hypoplastic left heart syndrome (HLHS), Shone complex, ostium secundum atrial septal defects, atrial fibrillation, and dilated cardiomyopathy (17–19). Depending on the cohort studied, MYH6 mutations are found in up to 10% of HLHS cases and predict poor clinical outcomes (19–22). BAV frequently co-occurs with HLHS and co-segregates with HLHS in families. BAV and HLHS are also caused by similar chromosomal abnormalities (Turner, Velocardiofacial, and Down syndromes). Therefore, MYH6 mutations most likely cause BAV in the context of other left-sided congenital lesions and may modify the clinical prognosis of BAV patients (19).

SMAD6 encodes an inhibitor of bone morphogenic protein signaling and is highly expressed in cardiac valves and the aortic root (23,24). SMAD6 mutations have been described in 3-9% of individuals with BAV and TAA, including predicted loss of function mutations that are similar to the frameshifts and indel that we identified in 3 EBAV probands (25,26). The SMAD6 mutant phenotype frequently includes other left ventricular obstructive lesions, such as coarctation and HLHS, as well as conotruncal defects, and these variable phenotypes are recapitulated in mice with knockout of the SMAD6 analog Madh6 (23,26–28). There is some evidence that SMAD6 functions downstream of NOTCH1 signaling to regulate endothelial cell motility and adhesion (28). Notably, other candidate genes (JAG1, MIB1) also participate in this pathway. Loss of this function during cardiac development may predispose to BAV (28).

Mutations of fibrillin and collagen genes can cause overlap syndromes with features of Ehlers-Danlos syndrome (EDS) hypermobility, cardiac valvulopathies, or TAD. We identified recurrent rare and predicted pathogenic variants of FBN2 and type 1 and type 5 collagens in the EBAV cohort. COL1A1 or COL1A2 mutations are the major cause of osteogenesis imperfecta (OI) with variable aortic and mitral valve disease and features of EDS (29,30). Up to half of COL1 mutation carriers develop cardiac complications, primarily aortic regurgitation due to a dysplastic or bicuspid aortic valve (31). COL1 mutations also predispose to sudden death due to TAD or intracranial aneurysms and dissections (32). Type 1 and Type 5 collagens are highly expressed in the developing aortic valve and in the adventitia of blood vessels (33). Type 5 collagens form heterotypic fibrils with type 1 collagens and are required for the assembly of type 1 collagen fibers (33,34). Approximately 50% of classical EDS cases are caused by mutations of COL5A1 or COL5A2 (35,36). COL5A1 has been implicated as the causal gene in several families with EDS features who also present due to arterial dissections (32,37–39). COL5A2 mutations and copy number variants are enriched in cohorts with cervical arterial dissections and may also predispose to TAD (40–43). Notably, low penetrance BAV has been described as a feature of classical EDS, and rare mutations of COL5A1 and COL5A2 have been observed in other BAV cohorts (44–46). Mutations of FBN2 encoding fibrillin 2, which cause congenital contractural arachnodactyly with incompletely penetrant TAD, were also identified as a potential modifier for BAV disease based on low penetrance variants in a younger subgroup of cases with more aggressive complications (45). We hypothesize that deleterious variants of extracellular matrix components such as collagens or fibrillins probably do not cause BAV but may accelerate the onset and severity of BAV disease, explaining their enrichment in EBAV cases.

Mutations of KCNH2 cause several types of cardiac conduction abnormalities predisposing to arrhythmias and sudden cardiac death, including Long QT syndrome, Romano-Ward syndrome, Short QT syndrome, Brugada syndrome, and familial atrial fibrillation (47). KCNH2 is one of several genes with recurrent missense and loss of function mutations in the EBAV cohort that encode components of cardiac myocyte ion channels, along with HCN4, KCNQ1, SCN5A, SCN8A, and SCN10A. KCNQ1, SCN5A, SCN8A, and SCN10A mutations can also cause sudden cardiac death by disrupting potassium or sodium channel activities (48–50). HCN4 mutations are implicated in familial sinus bradycardia, left ventricular noncompaction cardiomyopathy (LVNC), and TAD (51–53). Intriguingly, LVNC was also identified in families with KCNH2, KCNQ1 and SCN5A mutations (52–55). The mechanistic connection between conduction abnormalities and LVNC in channelopathy mutants is not currently known, but abnormal cardiac and vascular smooth muscle contraction during heart morphogenesis may contribute to impaired ventricular remodeling (56). The prevalence of LVNC is generally increased in BAV cohorts, and mutation of KCNJ2, another potassium channel subunit, causes a syndromic presentation of BAV with extracardiac features in addition to arrhythmias (57). BAV and TAD also co-segregate with LVNC in some families with mutations in NKX2-5, TBX20 or MIB1, another Notch pathway regulator that was convincingly linked to BAV in recent studies and is recurrently mutated in the EBAV cohort (52,58,59). Our findings provide further evidence for a genetic link between cardiomyopathies, BAV, conduction abnormalities, and TAD that may contribute to early onset complications of BAV disease.

Heterozygous truncating mutations of PKP2 are a common cause of arrhythmogenic right ventricular cardiomyopathy (ARVC) but have also been linked to LVNC in rare cases (52,60,61). Two other genes with loss of function variants in EBAV probands, JUP and MYH7, are also moderately or definitively linked to LVNC (52,53). PKP2 and JUP encode components of the cardiac desmosome, but more broadly anchor an intercalated disk scaffold called the connexome, which regulates intracellular signaling in cardiomyocytes (62). Disruption of the connexome alters the activity of sodium channels including SCN5A and contributes to arrhythmias in ARVC patients with PKP2 or JUP mutations (61,62). Gene therapies that restore myocardial expression of JUP or PKP2 can rescue structural heart and conduction ARVC phenotypes in mice by preventing connexome attrition (63). Components of the connexome are required in second heart field precursors for normal heart development (61). It is not known if the molecular functions of the connexome are also required for outflow tract or aortic valve formation. However, the prevalence of BAV is not known to be enriched in ARVC patients.

KIF1A and SHANK3 are enriched in neural tissues and are associated with neurodevelopmental disorders such as autism, epilepsy, or spastic paraplegia (64). KIF1A encodes a kinesin-3 family microtubular motor required for organelle transport. SHANK3 encodes a scaffold protein that anchors actin filaments to the cell membrane and regulates cell motility (65). However, both genes are also highly expressed in the developing heart and have been implicated in cardiac malformations (66). Overexpression of the Drosophila *kif1A* ortholog results in a hypertrophic cardiomyopathy phenotype with cardiac valve malformations (67). Balanced translocations involving the KIF1A gene were observed in HLHS (67). Mutation of a mouse Kif1a paralog causes situs inversus with left-sided cardiac lesions including BAV (68). Mutations of SHANK3 cause Phelan-McDermid syndrome, which is characterized by a variable phenotype of neurodevelopmental abnormalities, syndromic facial features, and cardiac abnormalities including BAV (69).

## Conclusion

In whole exome sequences of 215 probands with early onset complications of BAV disease, we identified likely pathogenic variants of CHD genes in 82 EBAV families (38%), including predicted pathogenic mutations that are known to cause BAV and HTAD. We identified variants in 27 other candidate genes that are associated with dominant cardiac phenotypes, including loss of function mutations in *PKP2*, *MYH6*, *KCNH2, KIF1A*, and *KMT2D*. Predicted pathogenic variants in *COL1A2*, *COL5A1*, *FBN2*, *MIB1*, *MYH6*, and *NOTCH3* were also observed in a GenTAC BAV cohort and in cohorts with familial or early onset HTAD. The findings from this study corroborate our hypothesis that genetic variants promoting the early onset of BAV disease are distinct and overlap with genetic variants that cause other congenital left-sided cardiac lesions. Genetic testing for predictive variants in these candidate genes may eventually prove useful to identify patients who may benefit from more aggressive surveillance or therapies to prevent BAV complications.

## Limitations

Genetic and phenotypic data on many EBAV families were incomplete, and the sample size of the EBAV cohort was insufficient for subgroup analysis. These factors likely decreased penetrance estimates and reduced sensitivity to detect genotype-phenotype correlations. Some contributory genetic variants may have been filtered out when recurrent rare variants were prioritized during the analysis. The limited evidence for familial segregation suggests that some variants are unlikely to cause BAV but modify the onset of clinically apparent disease. Future functional studies may be needed to validate variant effects and define the predictive utility of genetic testing.

## Data Availability

All data produced in the present study are available upon reasonable request to the authors

## Acknowledgments

We thank Joana Castillo and Jacqueline Jennings for sample preparation, and Jeff Weiss at the University of Washington Center for Rare Disease Research for whole exome sequence data preparation and assistance with data analysis.

The EBAV Investigators are: Shaine A. Morris, Rita Milewski, Giuseppe Limongelli, Allesandro Della Corte, Laura Perrone, Yuli Y. Kim, Hector Michelena, Maria G. Andreassi, Arturo Evangelista, Denver Sallee, Anji Yetman, Kim McBride, Eduardo Bossone, Rodolfo Citro, Dawn S. Hui, Malenka M. Bissell, Andrea Ballotti, Ilenia Foffa, Margot De Marco, Anthony Caffarelli, Rita Weise, Julie DeBacker, Laura Muino Mosquera, Robbin Cohen, Laura Dos Subira, Justin T. Tretter, Anna Sabe Rotes, Martina Caiazza, Lamia Ait Ali, Francesca Pluchinotta, and Simon C. Body.

## Sources of Funding

This study was supported in part by R01HL137028 (SP), R21HL150383 (SP), R01HL114823 (SCB), and R21HL150373 (SCB).

## Notes

### Competing Interest Statement

The authors have declared no competing interest.

### Author Declarations

The Committee for the Protection of Human Subjects at the University of Texas Health Science Center at Houston gave ethical approval for this work.

