## Supplemental Data for "Whole Exome Sequencing Uncovers the Genetic Complexity of Bicuspid Aortic Valve in Families with Early Onset Complications"

**Supplemental Methods: Whole Exome Sequencing**

The UW-Center for Rare Disease Research (CRDR) has the technical staff to carry out all sample processing steps for second-generation sequencing, including DNA quality control/assurance, library construction, targeted, in-solution exome capture methods, DNA sequencer operation and maintenance, variant calling, data analysis, and IT support. The UW-CRDR centralizes all receipt, tracking, and quality control/assurance of DNA samples. Samples have a detailed sample manifest (i.e., identification number/code, sex, DNA concentration, barcode, extraction method). Initial QC entailed DNA quantification, sex typing, and molecular “fingerprinting” using a 63-SNP Open Array assay derived from a custom exome SNP set. Samples were failed if: (1) the total amount, concentration, or volume was too low; (2) the fingerprint assay produced poor genotype data or integrity of DNA; or (3) sex-typing was inconsistent with the sample manifest. Library construction and exome capture were automated (Perkin-Elmer Janus II) in 96-well plate format. 500ng of genomic DNA was subjected to a series of shotgun library construction steps, including fragmentation through acoustic sonication (Covaris), end-polishing and A-tailing, ligation of sequencing adaptors, and PCR amplification with dual 10bp barcodes for multiplexing. Libraries underwent exome capture using the Twist exome (36.5MB target) (Twist Biosciences). Prior to sequencing, the library concentration was determined by fluorometric assay and molecular weight distributions were verified on the Agilent Bioanalyzer (consistently 180 ± 15bp). Barcoded exome libraries were pooled using liquid handling robotics prior to loading. Massively parallel sequencing-by-synthesis with fluorescently labeled, reversibly terminating nucleotides was carried out on the NovaSeq sequencer. Variant detection and genotyping were performed using the Haplotype Caller (HC) tool from GATK (3.7). Variant data for each sample were formatted (variant call format [VCF]) as “raw” calls that contain individual genotype data for one or multiple samples and flagged using the filtration walker (GATK) to mark sites that were of lower quality or false positives [e.g., low quality scores (Q50), allelic imbalance (ABHet 0.75), long homopolymer runs (HRun> 4), and/or low quality by depth (QD < 5)].

| **BAV** | **HTAD** | **CHD** | **CHD** | **CHD** | **CHD** | **CHD** | **CHD** | **CHD** |
| --- | --- | --- | --- | --- | --- | --- | --- | --- |
| CELSR1* | ACTA2 | ABCC9 | CFC1 | FOXC1 | KAT6B | NIPBL | RANBP9 | TBX1 |
| GATA4 | ADAMTS10 | ACTB | CFL1 | FOXC2* | KDM6A | NOTCH2 | RBM10 | TBX2 |
| GATA5 | BGN | ACVR1 | CHD4* | FOXF1 | KDR | NPHP3* | RBPJ | TBX3 |
| GATA6 | CBS | ACVR2B | CHD7 | FOXH1 | KIF7 | NR1D2 | RECQL4 | TBX5 |
| MAT2A | COL3A1 | ADAM17 | CITED2 | FOXJ1 | KMT2D* | NR2F2 | RFX3 | TFAP2B |
| MATR3 | COL5A1* | ADAMTS10 | CREBBP | FOXP1 | KRAS | NRAS | RHOA | TP53 |
| MIB1* | COL5A2* | AKAP13 | CRELD1 | FUZ | LATS2 | NSD1 | RHOV | UBC |
| NKX2-5 | EFEMP2 | AKT1 | CSK | G6PC3 | LRP2* | OFD1 | RIT1 | UBR1 |
| NOTCH1* | FBN1 | APC | CTNNB1 | GALNT1 | LTBP1 | PABPC1 | ROCK1 | UBR4 |
| ROBO4 | FLNA | ARHGAP31 | DCHS1 | GDF1 | MAML1 | PBX1 | ROCK2 | VEGFA |
| SMAD6* | FOXE3 | ARHGEF11 | DDX10 | GJC1 | MAP2K1 | PCDHGA2 | ROR2 | YWHAB |
| TBX20 | HCN4* | ARID1B* | DDX3X | GLRX3 | MAP2K2 | PCSK5* | RPL5 | ZBTB14 |
|  | LOX | B3GALTL | DHCR7 | GRB2 | MAP3K5 | PCSK6 | RPS19 | ZEB2 |
|  | MED12 | B3GAT3 | DLL4 | HAND1 | MAPK1 | PDK1L1 | RPS27A | ZFPM1 |
|  | MYH11 | BBS2 | DNAH5 | HAND2 | MEGF8* | PKD1 | SALL1 | ZFPM2 |
|  | MYLK | BBS6 | DOCK6* | HDAC10 | MEIS2 | PRDM1 | SHOC2 | ZIC3 |
|  | PLOD1 | BCL9 | E2F6 | HES1 | MGRN1 | PRICKLE1 | SLIT3 |  |
|  | PRKG1 | BCOR | EFTUD2 | HEY2 | MINK1 | PRKACB | SMARCA4 |  |
|  | SKI | BMP1 | EHMT1 | HIC2 | MKKS | PRKCD | SMARCC1 |  |
|  | SLC2A10 | BMP4 | ELN | HOXA1 | MNDA | PRKCH | SMURF1 |  |
|  | SMAD2 | BMPR1A | EOGT | HRAS | MSX1 | PTK7 | SON |  |
|  | SMAD3 | BMPR2 | EP300 | HSP90AB1 | MYH6* | PTPN11 | SOS1 |  |
|  | SMAD4 | BRAF | EPHA2 | IGF1R | NAA15 | PYGL | SRC |  |
|  | TGFB2 | CAV1 | ESCO2 | ITPR3 | NCK1 | PYGO | SSH2 |  |
|  | TGFB3 | CBP | ETS1 | JAG1* | NEK8 | RAB23 | STRA6 |  |
|  | TGFBR1 | CCDC91 | EVC | JARID2 | NF1 | RAC1 | SUFU |  |
|  | TGFBR2 | CDH2 | EVC2* | JMJD6 | NFATC1 | RACK1 | TAB1 |  |
|  | THSD4 | CDK13 | FGF19 | KANSL1 | NGFR | RAF1 | TAB2 |  |
|  |  | CDK4 | FLT4* | KAT6A | NIFK | RAI1 | TBC1D32 |  |

**Table S1. Medium- to high-confidence BAV, HTAD, and CHD genes evaluated in EBAV cohort.**

| **Gene** | **Chr** | **BP** | **Ref > Alt** | **Amino Acid** | **Consequence** | **gnoMAD** |
| --- | --- | --- | --- | --- | --- | --- |
| CELSR1 | 22 | 46367090 | A > T | p.Val2703Glu | Missense | 51 |
| CELSR1 | 22 | 46411758 | G > A | p.Pro1538Leu | Missense | 2 |
| MIB1 | 18 | 21864630 | T > TC | p.Arg996ProfsTer5 | Frameshift | 0 |
| MIB1 | 18 | 21765808 | G > A | p.Arg89His | Missense | 14 |
| MIB1 | 18 | 21857207 | G > A | p.Gly915Arg | Missense | 27 |
| NOTCH1 | 9 | 136509896 | C > T | p.Gly936Ser | Missense | 14 |
| NOTCH1 | 9 | 136505319 | C > T | p.Gly1526Asp | Missense | 0 |
| ROBO4 | 11 | 124887023 | C > A | p.Glu797Ter | Stop Gained | 0 |
| SMAD6 | 15 | 66781184 | GC > G | p.Arg381GlyfsTer158 | Frameshift | 0 |
| SMAD6 | 15 | 66781118 | CG > C | p.Asp359ThrfsTer180 | Frameshift | 0 |
| SMAD6 | 15 | 66703331 | G > GGCGGCA | p.Ser27_Gly28dup | In frame Ins | 7 |
| SMAD6 | 15 | 66703830 | T > C | p.Leu191Pro | Missense | 5 |
| SMAD6 | 15 | 66711706 | G > A | p.Asp286Asn | Missense | 79 |
| ACTA2 | 10 | 88935264 | C > T | p.Asp365Asn | Missense | 4 |
| COL5A1 | 9 | 134765688 | G > A | p.Arg681His | Missense | 26 |
| COL5A1 | 9 | 134768446 | G > A | p.Gly757Ser | Missense | 7 |
| COL5A1 | 9 | 134802904 | C > T | p.Thr1008Met | Missense | 132 |
| COL5A1 | 9 | 134750794 | G > A | p.Arg525Gln | Missense | 61 |
| COL5A1 | 9 | 134750857 | C > T | p.Ala546Val | Missense | 176 |
| COL5A1 | 9 | 134802895 | C > T | p.Thr1005Met | Missense | 157 |
| COL5A1 | 9 | 134834992 | G > A | p.Gly1720Ser | Missense | 2 |
| FBN1 | 15 | 48503897 | C > T | p.Gly668Asp | Missense | 0 |
| FBN2 | 5 | 128277982 | G > A | p.Pro2457Ser | Missense | 2 |
| FBN2 | 5 | 128393366 | C > T | p.Glu412Lys | Missense | 1 |
| FBN2 | 5 | 128301501 | T > C | p.Glu1976Gly | Missense | 0 |
| FBN2 | 5 | 128350005 | T > C | p.Asp938Gly | Missense | 0 |
| FBN2 | 5 | 128328713 | T > C | p.Asp1485Gly | Missense | 437 |
| FBN2 | 5 | 128328749 | C > T | p.Arg1473His | Missense | 76 |
| FLNA | X | 154354973 | G > A | p.Thr1690Met | Missense | 12 |
| FOXE3 | 1 | 47416464 | Deletion | p.Thr54AlafsTer217 | Frameshift | 0 |
| HCN4 | 15 | 73329671 | T > C | p.Ser498Gly | Missense | 0 |
| MYH11 | 16 | 15735546 | C > A | p.Ala1116Asp | Missense | 0 |
| SMAD4 | 18 | 51058457 | Deletion | Frameshift | Structural | 152 |
| THSD4 | 15 | 71748523 | C > A | p.Pro782Thr | Missense | 26 |

**Table S2. BAV and HTAD Gene Mutations in EBAV Cohort**

| **Gene** | **Chr** | **BP** | **Ref > Alt** | **Amino Acid** | **Consequence** | **gnoMAD** |
| --- | --- | --- | --- | --- | --- | --- |
| COL1A1 | 17 | 50189879 | G > A | p.Arg865Cys | Missense | 12 |
| COL1A1 | 17 | 50187967 | C > T | p.Arg1093His | Missense | 111 |
| COL1A1 | 17 | 50188764 | C > T | p.Arg1026Gln | Missense | 8 |
| COL1A1 | 17 | 50191799 | C > T | p.Asp706Asn | Missense | 125 |
| COL1A2 | 7 | 94427849 | C > T | p.Arg1164Cys | Missense | 13 |
| COL1A2 | 7 | 94427852 | G > A | p.Asp1165Asn | Missense | 6 |
| COL1A2 | 7 | 94409359 | G > C | p.Gly277Ala | Missense | 61 |
| COL1A2 | 7 | 94405676 | G > A | p.Ala164Thr | Missense | 9 |
| COL1A2 | 7 | 94420275 | C > T | p.Arg708Trp | Missense | 11 |
| COL1A2 | 7 | 94423018 | G > A | p.Arg822His | Missense | 78 |
| COL5A3 | 19 | 9978592 | TG > T | p.Pro1000GlnfsTer144 | Frameshift | 33 |
| COL5A3 | 19 | 9996616 | G > A | p.Pro446Leu | Missense | 29 |
| COL5A3 | 19 | 9989359 | G > T | p.Pro685Gln | Missense | 33 |
| COL5A3 | 19 | 9992873 | C > T | p.Arg601Gln | Missense | 9 |
| EVC2 | 4 | 5640789 | G > A | p.Arg399Ter | Stop Gained | 45 |
| EVC2 | 4 | 5622564 | A > G | p.Leu825Pro | Missense | 2 |
| EVC2 | 4 | 5625864 | C > T | p.Arg644Gln | Missense | 54 |
| FLNC | 7 | 128853606 | G > A | p.Asp2116Asn | Missense | 2 |
| FLNC | 7 | 128837539 | T > C | p.Tyr281His | Missense | 0 |
| FLNC | 7 | 128847780 | A > G | p.Arg1458Gly | Missense | 2 |
| FLNC | 7 | 128855291 | C > T | p.Arg2410Cys | Missense | 23 |
| JAG1 | 20 | 10645364 | C > T | p.Cys702Tyr | Missense | 0 |
| JAG1 | 20 | 10648023 | C > T | p.Glu553Lys | Missense | 5 |
| JAG1 | 20 | 10649549 | C > T | p.Gly441Ser | Missense | 44 |
| JAG1 | 20 | 10658641 | G > A | p.Thr174Met | Missense | 59 |
| KCNH2 | 7 | 5640789 | G > A |  | Frameshift | 0 |
| KCNH2 | 7 | 150955469 | CCT > C | p.Ala13SerfsTer34 | Frameshift | 8 |
| KCNH2 | 7 | 150957458 | C > A | p.Asp321Tyr | Missense | 2 |
| KCNH2 | 7 | 150951790 | C > T | p.Val535Met | Missense | 1 |
| KCNH2 | 7 | 150945376 | G > A | p.Pro1157Ser | Missense | 15 |
| KCNH2 | 7 | 150955440 | G > A | p.Arg22Trp | Missense | 5 |
| KIF1A | 2 | 240784988 | CCTT > C | p.Lys240del | In frame Del | 3 |
| KIF1A | 2 | 240725305 | G > C | p.Leu1408Val | Missense | 1 |
| KIF1A | 2 | 240763253 | G > A | p.Thr621Met | Missense | 88 |
| KIF1A* | 2 | 240724038 | T > G |  | Splicing | 0 |
| MEGF8 | 19 | 42336316 | T > C | p.Val405Ala | Missense | 15 |
| MEGF8 | 19 | 42358179 | A > C | p.Thr1683Pro | Missense | 1 |
| MEGF8 | 19 | 42356983 | T > C |  | Splicing | 8 |
| MEGF8 | 19 | 42370784 | C > T | p.Arg2Ter | Stop Gained | 0 |
| MYH6 | 14 | 23397212 | G > A | p.His670Tyr | Missense | 2 |
| MYH6 | 14 | 23388189 | G > A | p.Ala1442Val | Missense | 23 |
| MYH6 | 14 | 23393454 | T > C | p.Lys998Arg | Missense | 3 |
| MYH6 | 14 | 23393809 | C > G | p.Glu929Gln | Missense | 1 |
| MYH6 | 14 | 23388186 | G > T | p.Ala1443Asp | Missense | 327 |
| MYH6 | 14 | 23400708 | C > T |  | Splicing | 1 |
| MYH6 | 14 | 23393709 | G > A | p.Thr962Ile | Missense | 1 |
| NOTCH3 | 19 | 15165433 | G > A | p.Ala1917Val | Missense | 0 |
| NOTCH3 | 19 | 15181576 | C > A | p.Ser931Ile | Missense | 1 |
| NOTCH3 | 19 | 15191496 | C > G | p.Val322Leu | Missense | 1 |
| NOTCH3 | 19 | 15162478 | A > G | p.Met1967Thr | Missense | 0 |
| PKP2 | 12 | 32802499 | GGGTGT > G | p.His733ProfsTer8 | Frameshift | 113 |
| PKP2 | 12 | 32879021 | G > A | p.Arg79Ter | Stop Gained | 22 |
| PKP2 | 12 | 32896563 | G > A | p.Gln57Ter | Stop Gained | 0 |
| PTPN13 | 4 | 86758292 | TCAC > T | p.Pro1087del | In frame Del | 61 |
| PTPN13 | 4 | 86811047 | T > C | p.Phe2439Ser | Missense | 0 |
| PTPN13 | 4 | 86734792 | G > C | p.Glu690Gln | Missense | 1 |
| PTPN13 | 4 | 86807733 | A > G | p.Met2312Val | Missense | 9 |
| SCN10A | 3 | 38728868 | T > G | p.Thr772Pro | Missense | 0 |
| SCN10A | 3 | 38702079 | C > T | p.Val1473Met | Missense | 30 |
| SCN10A | 3 | 38792159 | C > T | p.Val94Met | Missense | 0 |
| SHANK3 | 22 | 50675096 | C > T | p.Arg38Cys | Missense | 3 |
| SHANK3 | 22 | 50679030 | C > A | p.Asp204Glu | Missense | 0 |
| SHANK3 | 22 | 50694903 | G > A | p.Ala387Thr | Missense | 0 |
| SHANK3 | 22 | 50697269 | T > G |  | Splicing | 0 |
| TPTE2 | 13 | 19464463 | G > A | p.Pro245Leu | Missense | 1 |
| TPTE2 | 13 | 19450090 | C > T | p.Cys320Tyr | Missense | 26 |
| TPTE2 | 13 | 19465494 | T > A | p.Arg195Ter | Stop Gained | 2 |

**Table S3. CHD gene mutations (>=3 families) in EBAV Cohort**

*De novo variant

| **Gene** | **Chr** | **BP** | **Ref > Alt** | **Amino Acid** | **Consequence** | **gnoMAD** |
| --- | --- | --- | --- | --- | --- | --- |
| APC | 5 | 112839733 | C > G | p.Thr1380Ser | Missense | 2 |
| CHD4 | 12 | 6583081 | C > T | p.Asp1365Asn | Missense | 6 |
| CHD4 | 12 | 6593463 | A > G | p.Phe823Leu | Missense | 0 |
| DOCK6 | 19 | 11243094 | G > A | p.Pro482Leu | Missense | 54 |
| DOCK6 | 19 | 11235638 | G > T | p.Tyr838Ter | Stop Gained | 1 |
| FLT4 | 5 | 180628915 | T > A | p.Lys357Met | Missense | 3 |
| FLT4 | 5 | 180619769 | C > T | p.Gly848Glu | Missense | 33 |
| FOXC2 | 16 | 86567754 | C > T | p.Pro140Leu | Missense | 0 |
| FOXC2 | 16 | 86567775 | C > T | p.Thr147Ile | Missense | 0 |
| FOXJ1 | 17 | 76137706 | G > C | p.Leu305Val | Missense | 0 |
| FUZ | 19 | 49812717 | C > T | p.Gly44Asp | Missense | 0 |
| GDF1 | 19 | 18868856 | G > A | p.Ala287Val | Missense | 2 |
| HRAS | 11 | 533536 | G > A | p.Arg123Cys | Missense | 28 |
| JUP | 17 | 41765060 | AT > A | p.Ile306SerfsTer56 | Frameshift | 26 |
| JUP | 17 | 41764938 | C > T | p.Ala347Thr | Missense | 40 |
| KMT2D | 12 | 49043927 | GCTT > G | p.Lys1753del | In frame Del | 25 |
| KMT2D | 12 | 49043887 | A > T | p.Met1767Lys | Missense | 1 |
| LRP1 | 12 | 57208809 | G > A | p.Trp4046Ter | Stop Gained | 0 |
| LRP1 | 12 | 57199905 | T > G | p.Asn3298Lys | Missense | 5 |
| LRP2 | 2 | 169206749 | C > T | p.Arg2324Lys | Missense | 179 |
| LRP2 | 2 | 169212118 | C > T | p.Ala2044Thr | Missense | 220 |
| PCSK5 | 9 | 76332527 | A > AGG | p.Arg1530GlyfsTer17 | Frameshift | 2 |
| PCSK5 | 9 | 75891353 | G > A | p.Gly58Arg | Missense | 19 |
| PKD1 | 16 | 2091559 | G > A | p.Pro3859Leu | Missense | 2 |
| PTK7 | 6 | 43129171 | C > T | p.Arg100Trp | Missense | 13 |
| ROR2 | 9 | 91757318 | G > C | p.Asn139Lys | Missense | 0 |
| SCN8A* | 12 | 51806641 | CCT > C | p.Pro1719ArgfsTer6 | Frameshift | 1 |
| SMARCA4 | 19 | 11034168 | G > A | p.Ala1307Thr | Missense | 2 |
| TBX1 | 22 | 19763243 | G > A | p.Arg138Gln | Missense | 1 |
| ZEB2 | 2 | 144517323 | G > A | p.Pro10Ser | Missense | 2 |

**Table S4. CHD gene mutations (<3 families) in EBAV Cohort**

*De novo variant

| **Gene** | **Variant** | **Family** | **Affected relatives with variant** | **Affected relatives without variant** | **Unaffected relatives with variant** | **Unaffected relatives without variant** | **Important Notes** |
| --- | --- | --- | --- | --- | --- | --- | --- |
| COL5A1 | Missense | BAV125 | 1 | 0 | 1 | 0 |  |
| KCNH2 | Missense | BAV125 | 1 | 0 | 0 | 1 |  |
| COL1A2 | Missense | BAV185 | 1 | 0 | 2 | 0 |  |
| FBN2 | Missense | BAV254 | 1 | 0 | 0 | 2 |  |
| COL1A1 | Missense | BAV271 | 1 | 0 | 1 | 1 |  |
| ROBO4 | Stop Gained | BAV277 | 1 | 0 | 1 | 2 |  |
| COL1A2 | Missense | BAV287 | 1 | 0 | 0 | 1 |  |
| SMAD6 | Missense | BAV305 | 0 | 1 | 2 | 2 | Variant not in proband |
| NOTCH3 | Missense | BAV308 | 2 | 0 | 0 | 1 |  |
| CHD4 | Missense | BAV328 | 1 | 0 | 0 | 1 |  |
| SMAD6 | In frame Ins | BAV336 | 0 | 1 | 1 | 1 | Variant not in proband |
| COL5A1 | Missense | BAV338 | 1 | 0 | 1 | 1 |  |
| PKP2 | Frameshift | BAV338 | 1 | 0 | 1 | 1 |  |
| KCNH2 | Missense | BAV375 | 1 | 0 | 0 | 1 |  |
| COL1A2 | Missense | BAV387 | 1 | 0 | 2 | 1 |  |
| SCN8A | Frameshift | BAV433 | 1 | 0 | 0 | 4 |  |
| ADCY7 | Frameshift | BAV438 | 1 | 0 | 0 | 1 |  |
| COL5A3 | Missense | BAV502 | 1 | 0 | 1 | 1 |  |
| CACNA1H | Missense | BAV522 | 3 | 0 | 0 | 1 |  |
| MYH6 | Missense | BAV522 | 1 | 2 | 0 | 1 |  |
| JAG1 | Missense | BAV543 | 1 | 0 | 1 | 1 |  |
| KIF1A | Splicing | BAV546 | 1 | 0 | 0 | 5 |  |
| SHANK3 | Missense | BAV546 | 1 | 0 | 0 | 5 |  |
| PTPN13 | Missense | BAV787 | 1 | 0 | 0 | 2 |  |
| JAG1 | Missense | BAV817 | 1 | 0 | 1 | 1 |  |
| FBN1 | Missense | BAV894 | 1 | 0 | 0 | 1 |  |
| PTPN13 | Missense | BAV903 | 1 | 0 | 0 | 1 |  |
| JUP | Missense | BAV990 | 2 | 0 | 0 | 0 |  |

**Table S5. Table Segregation of Rare Variants in EBAV Cohort.**

| **Family** | **Variants** | **Valve Function** | **Aortic and Valvular Interventions** | **Aorta and Other Congenital Anomalies** |
| --- | --- | --- | --- | --- |
| BAV002 | MYH6 missense | AR |  |  |
| BAV036 | COL1A2 missense, MEGF8 stop gained | AR | Aortic valve repair and ascending aortic replacement | Large ascending aneurysm |
| BAV048 | KMT2D indel | AR |  | Mitral valve prolapse |
| BAV049 | ACTA2 missense | AR | Ascending aortic replacement | Large ascending aneurysm |
| BAV065 | COL5A1 missense | AR, AS | Aortic valve replacement, coarctation repair | Root and ascending aneurysm, coarctation, VSD |
| BAV090 | CELSR1 missense, NOTCH1 missense, SMAD6 frameshift |  | Ascending aortic replacement | Large root and ascending aneurysm |
| BAV120 | COL1A2 missense | AR |  | Ehlers-Danlos syndrome |
| BAV125 | COL5A1 missense, KCNH2 missense |  |  |  |
| BAV128 | SMAD6 frameshift | AS |  | Ascending aneurysm |
| BAV148 | MEGF8 splice, PTPN13 indel, COL1A1 missense, FLNA missense | AS |  |  |
| BAV162 | NPHP3 missense, FOXE3 frameshift | AR |  | Mitral valve prolapse, Root aneurysm |
| BAV185 | COL1A2 missense |  | Aortic valve replacement |  |
| BAV198 | FBN2 missense | AR |  |  |
| BAV215 | FLNC missense, KCNH2 missense |  | Aortic valve repair |  |
| BAV254 | FBN2 missense | AR |  | Large ascending aneurysm |
| BAV263 | CELSR1 missense | AS, AR | Aortic valve replacement (Ross) |  |
| BAV271 | COL1A1 missense | AR |  | Tricuspid valve prolapse |
| BAV277 | ROBO4 stop gained | AR |  | VSD |
| BAV287 | COL1A2 missense | AS |  | Coarctation, ASD |
| BAV305 | SMAD6 missense | AS, AR | Aortic valve repair and ascending aortic replacement | Large ascending aneurysm |
| BAV308 | NOTCH3 missense | AS, AR |  |  |
| BAV315 | CHD4 missense, COL1A1 missense, CACNA1H splice | AS | Aortic root replacement and aortic valve replacement | Large root aneurysm |
| BAV326 | MIB1 frameshift | AS, AR | Aortic valve replacement (Ross), ascending aortic replacement |  |
| BAV328 | CHD4 missense | AS | Coarctation repair | Coarctation, Dextrocardia, Mitral valve prolapse |
| BAV330 | LRP2 splice, MYH6 missense | AR |  |  |
| BAV334 | EVC2 missense, SHANK3 missense | AS |  | Root aneurysm |
| BAV336 | SMAD6 inframe insertion | AR |  | Unicommisural AV |
| BAV337 | FBN2 missense, LRP1 missense, SMAD4 splice | AS |  |  |
| BAV338 | COL5A1 missense, PKP2 frameshift | AS |  |  |
| BAV348 | LRP1 stop gained | AS | Aortic valvuloplasty | Kawasaki disease with coronary aneurysms |
| BAV375 | KCNH2 missense | AR |  | Junctional ectopic tachycardia and non-sustained VT |
| BAV387 | COL1A2 missense | AS | Aortic valvuloplasty |  |
| BAV488 | EVC2 stop gained, NPHP3 missense | AR | Aortic valve replacement | Large ascending aneurysm, CHARGE syndrome |
| BAV494 | MYH6 splice | AS |  | Coarctation |
| BAV522 | MYH6 missense | AS |  |  |
| BAV528 | CACNA1H missense, FBN2 missense | AS, AR |  | Unicommisural AV, Large ascending aneurysm |
| BAV533 | SCN10A missense | AS |  | Ascending aneurysm |
| BAV543 | JAG1 missense | AS |  | VSD, ASD, PDA, Large ascending aneurysm |
| BAV546 | KIF1A splice, SHANK3 missense | AR |  | Coarctation, VSD |
| BAV581 | NOTCH1 missense | AS, AR | Aortic valve replacement, ascending aortic replacement |  |
| BAV583 | KIF1A missense |  | Ascending aortic replacement |  |
| BAV780 | MEGF8 missense | AR | Coarctation repair | Coarctation |
| BAV787 | PTPN13 missense | AS, AR | Aortic valve replacement, aortic root replacement, ascending aortic replacement | Large ascending and root aneurysm |
| BAV816 | MIB1 missense | AR |  |  |
| BAV817 | JAG1 missense | AR | Coarctation repair | Sievers 0 A-P, Coarctation |
| BAV827 | COL5A1 missense | AS, AR |  |  |
| BAV846 | KCNH2 frameshift | AR |  | Large ascending aneurysm, history of congestive heart failure after elective surgery in two relatives with BAV |
| BAV855 | COL5A3 frameshift, LRP2 missense, NOTCH3 missense, PKD1 missense | AS, AR | Aortic valve replacement, ascending aortic replacement | Large ascending aneurysm |
| BAV874 | MYH6 missense | AR | Ascending, arch, and descending aortic replacement | Sievers 0 A-P valve, Large ascending, arch, and proximal descending aneurysm |
| BAV877 | FLNC missense, LRP2 missense |  |  | Mitral valve prolapse, Ehlers Danlos syndrome |
| BAV892 | THSD4 missense | AR |  | Ascending aneurysm, Mitral valve prolapse, Ehlers-Danlos syndrome |
| BAV914 | HCN4 missense, HRAS missense, SMAD6 missense | AR | Aortic valve replacement, ascending aortic replacement, mitral valve replacement | Mitral valve prolapse, Giant anterior mitral leaflet |
| BAV917 | FLT4 missense |  |  | Sievers 0 A-P valve, Large ascending aneurysm |
| BAV927 | MIB1 missense, NOTCH3 missense |  |  |  |
| BAV934 | KMT2D missense, SCN10A missense | AR | Ascending aortic replacement | Large ascending aneurysm |
| BAV946 | MEGF8 missense |  |  | Coarctation, ASD, VSD |
| BAV950 | COL5A1 missense, PKP2 stop gained | AS, AR | Aortic valve replacement and aortic root replacement |  |
| BAV951 | FLNC missense | AS, AR | Aortic valve replacement and aortic root replacement | Coarctation |
| BAV952 | PKP2 stop gained | AR |  | Coarctation |
| BAV958 | JAG1 missense, LRP2 missense | AS | Aortic valve repair, aortic root replacement |  |
| BAV961 | KIF1A missense, MYH11 missense |  |  | VSD |
| BAV962 | JAG1 missense | AS, AR | Aortic valve replacement |  |
| BAV963 | COL5A1 missense, KCNH2 missense | AS |  | VSD |
| BAV968 | COL5A1 missense, FBN2 missense | AR | Aortic valve replacement | Coarctation |
| BAV975 | FLNC missense, FLT4 missense, PTPN13 missense, TPTE2 stop gained | AR | Aortic valve replacement, ascending aortic replacement | Large ascending aneurysm |
| BAV978 | COL1A1 missense | AR |  |  |
| BAV980 | COL1A2 missense, DOCK6 stop gained | AS, AR |  |  |
| BAV982 | FOXC2 missense x2, FOXJ1 missense | AR | VSD patch closure | Coarctation, VSD |
| BAV985 | SCN10A missense | AS |  | Large ascending and root aneurysm |
| BAV987 | FBN2 missense | AR | VSD patch closure | Coarctation, VSD |

**Table S6. Phenotypes of EBAV Probands With Rare Variants**
